# Modeling and reviewing analysis of the COVID-19 epidemic in Algeria with diagnostic shadow

**DOI:** 10.1101/2021.06.09.21258668

**Authors:** Jiwei Jia, Siyu Liu, Yawen Liu, Ruitong Shan, Khaled Zennir, Ran Zhang

## Abstract

In this paper, we formulate a special epidemic dynamic model to describe the transmission of COVID-19 in Algeria. We derive the threshold parameter control reproduction number 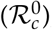, and present the effective control reproduction number (*R*_*c*_(*t*)) as a real-time index for evaluating the epidemic under different control strategies. Due to the limitation of the reported data, we redefine the number of accumulative confirmed cases with diagnostic shadow and then use the processed data to do the optimal numerical simulations. According to the control measures, we divide the whole research period into six stages. And then the corresponding medical resource estimations and the average effective control reproduction numbers for each stage are given. Meanwhile, we use the parameter values which are obtained from the optimal numerical simulations to forecast the whole epidemic tendency under different control strategies.

## 1 Introduction

Plague appears everywhere in human history and it always brings grave misery into society. In another word, the struggling against diseases play an important role in the establishment of mankind’s civilization. With the fast development of medical and pharmaceutical industry, some lethal infectious diseases can be cured completely, even the smallpox which plagued mankind for server centuries has been vanished. In recent years, there are small outbreaks in some areas all over the world which are not causing serious global outbreak. Nowadays, people think that we have already built up strong health system and developed high level of medical care.

However, at the end of 2019, everything changed. The first confirmed COVID-19 case was reported in Wuhan City, China [1]. At that time, because of the cognition limitation, it was called as an unknown infectious disease. The clinical symptoms are very similar with viral pneumonia [2, 3]. Within one month, it spreads rapidly in Wuhan City and causes many death. The serious anomaly quickly attracted the attention of Chinese government, strict isolation and lockdown strategies were implemented immediately [4]. The warning form China does not get enough attention by other countries. After a short period, the disease spreads all over the world, it has become a global outbreak. People have to stop nearly all kinds of human activities, due to the sudden pandemic causes by the COVID-19, which is an infectious disease of coronavirus.

People have to face the new infectious disease, it seems that the health system and quick drug research which we are proud of are not as effective as before. Unlike other plague happened in the past years, the COVID-19 has many different characteristics. The spread ability of the COVID-19 is really strong, the 2019-nCoV trimeric spike protein binds at least 10 times more tightly than the corresponding spike protein of SARS-CoV to their common host cell receptor [5]. In the initial stage, researchers believe that it is a kind of viral pneumonia, with more and more clinical cases, the lung is not the only damaged organ [6, 7]. According to the clinical studies, the COVID-19 attacks various organs of the body, including cardiac, gastrointestinal, hepatic, renal, neurologic, olfactory, gustatory, ocular, cutaneous and haematologic symptoms [8, 9]. The virus is very insidious, the incubation period is very complex and flexible. It may be latent in human body for several days or even several weeks, and more than half of people with positive nucleic acid test don’t show any typical clinical symptoms at all [10]. But the asymptomatic patients also have the ability to spread the disease [11]. A more severe problem is that, the structure of SARS-CoV-2 is enveloped, positive-sense single-stranded virus ((+)ssRNA virus) [12], the virus mutates in very fast speed [13], it makes great difficult in the developing of specific medicine, even worse, the vaccine may not be effective enough forever.

With the great challenge, people seem to come back to the Middle Ages, the most effective strategy is to protect the susceptible population. Social activities will increase the risk of exposure to SARS-CoV-2 and almost all countries published a series of lockdown strategies to prevent the disease from rapidly spreading. At first, in the view of traditional epidemiological experience, we just need to keep isolation for a few incubation period, but the variety incubation period and asymptomatic patient who are infectious make the lockdown strategy last for a long time. During the epidemic prevention period, it requires continuous cost of huge human and material resources. Basic medical facilities and financial expenditure of government are the most important parts in the control of the COVID-19. Long time lockdown strategy makes the unemployment rate higher. The COVID-19 causes a series of social problems and damages the economic system badly [14, 15]. These problems are particularly acute in developing countries. Whether the medical or economy resources are sufficient in developing countries, and the basic health system is weak or even non-existent there [16,17]. Now, before the vaccines come into use worldwide, people are trying to keep the balance between epidemic control and society development.

Many research appear about different aspects of the COVID-19 epidemic. Dynamical modeling is an effective mathematical tool for describing the spreading procedure of epidemic. Various dynamical model are proposed for modeling the COVID-19 transmission by many scholars. A model involving the interactive effect of various factors with delay effect for the COVID-19 transmission is proposed in [18]. A modified SEIR-type model is established in [19] with the effect of quarantine strategy taken into account, the estimated medical resource and the effect of vaccine are also analyzed. An impulsive dynamical model is employed to present the impact of imported population on the epidemic control in [20]. The effect of age-specific vaccination strategies, considering the age structure and social contact patterns for different age groups for each of different countries, is evaluated by an SEIR model in [21]. A deterministic model is developed to study the transmission dynamics of the COVID-19 with immigrant and local susceptible in [22], the existence of globally stable disease-free equilibrium point is also analyzed.

In this paper, we mainly focus on the epidemic control in Algeria which is a north African country, and the first COVID-19 confirmed case was reported on 25 Feb. 2020. The situation there is not optimistic, at the beginning of the outbreak, the average COVID-19 induced death rate in Algeria is about 15.7% until the middle of April, which is almost the highest in the world [23]. And by the end of November, the death rate caused by COVID-19 is as high as 3.06%, which is still higher than the average level around the world [24]. From the first confirmed case, the total trend of the new confirmed case goes high continuously. There are some studies on the epidemic of the COVID-19 in Algeria. In [25], the authors discuss the psychological impact of confinement, the results show that there is a significant change in people’s daily life. The control measures of the COVID-19 has been carried out since February. Algeria government perform neither the strict lock-down strategy in China, nor the herd immunity strategy such as Sweden, they introduce partial “confinement” measures in the most affected areas and most people are being advised to self-isolate in their houses to reduce social activities [26]. With the epidemic of the COVID-19 in Algeria getting better, the government reopen the country gradually.

In order to study the effectiveness of the control strategies and the regulation of spreading, in Section 2 we establish epidemic model to investigate the disease transmission process. Because the basic medical system is not very complete in Algeria, to have more realistic daily confirmed data, we introduce diagnostic shadow calculation into the data pre-processing in Section 3. According to the different control strategies, we divide the whole research process into six stages. The details and the algorithm of medical resource are presented in Section 3. We provide all the results of numerical simulations in Section 4 and conclude with a summary in Section 5. Our theoretical results and numerical simulations will help to have better understand of COVID-19 in Algeria.

## 2 Dynamical Model

In this section, according to the control strategy performed in Algeria, we establish the corresponding dynamical model to describe the development of the epidemic. Algeria is the largest country in the north of Africa. The economic level ranks top in Africa, the medical policy adopted there is free for the inhabitant. However, the medical level there is lag behind relatively to the developed countries and the primary health care facilities in some areas are scarce. The daily reported confirmed data does not distinguish the asymptomatic and symptomatic infected patients, so we only adopt class I to describe the infected population. The basic frame we choose in this paper is an SEIR model as well as there is a significant incubation period of COVID-19. And the control measures taken in Algeria is relative loose in general, so we cancel the group of lockdown strategy for susceptible population. Since the partial confinement measures are not compulsive, the restrictions on the movement of population is limited. Here we adopt home quarantine and hospitalization instead of the strict isolation group.

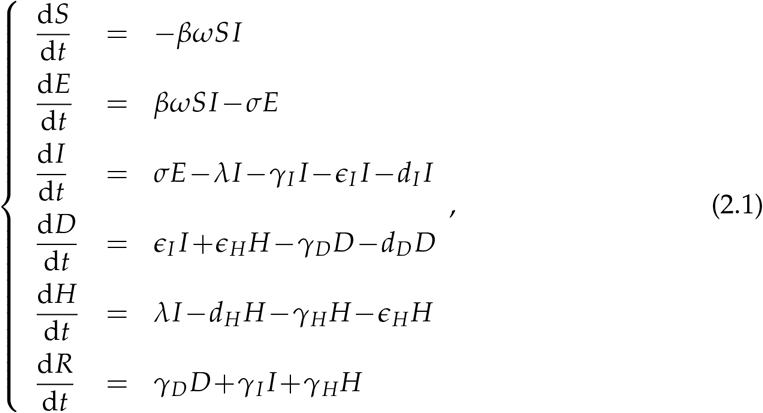

where *S*(*t*),*E*(*t*), *I*(*t*), *R*(*t*), *D*(*t*) and *H*(*t*) denote the individuals who are susceptible, exposed, infectious, recovered, hospitalization and home quarantined at time *t*, respectively. Some studies show that, during the late incubation period, the patients may infect the susceptible people [27]. Considering that, *E*(*t*) represents the compartment of low-level virus carriers, which cannot infect others. For the home quarantined *H*(*t*) and hospitalization *D*(*t*) compartments, we assume that they do not contact the susceptible individuals. The symptoms of home quarantined group are more mild than class *D*(*t*), totally. The relationship can be reflected from the parameters *γ*_*D*_ and *γ*_*H*_. Although some people will have slight symptoms at the early infected stage, but they may get worse suddenly. It is related to the autoimmune disorders and cytokine storm [28]. At this time, hospital treatment is necessary, we use parameter *ϵ*_*H*_ to describe the process.

Notice that, the disease-related death rate is very high in Algeria, it indicates that there are many undetected infected people. We introduce diagnostic shadow method to overcome the difficulty in the numerical simulations and the natural birth and death rate are ignored in the short-term model to highlight the actual level of disease-related death rate of class *I*(*d*_*I*_), *D*(*d*_*D*_) and *H*(*d*_*H*_). Definition of other parameters are explained as follows. The contact rate and transition rate of exposed to infected group are denoted as *β* and *σ*, respectively. The control strategies decrease the contact frequency of people and stop the disease spread to some extent, we define a piecewise-constant parameter *ω* to describe it and name it as the average level of social activity. The transition rate of infected population to home quarantined population and hospitalization are shown as *λ* and *ϵ*_*I*_. Parameter *γ*_*I*_, *γ*_*H*_ and *γ*_*D*_ represent the recovery rate of infected, hospitalization and home quarantined class, respectively. Totally, the tendency of disease symptom is weaken, so in each stage, the recovery rate of people is different and we estimate them in the numerical simulations.

### 2.1 Analysis of the control reproduction number

The epidemic model for the partial confinement strategy has been established, then we will discuss the important index to evaluate the control of the disease. The next generation matrix approach is applied to calculate the control reproduction number, the detail form is as follow:

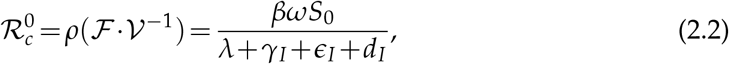

where *ρ* represents the spectral radius and

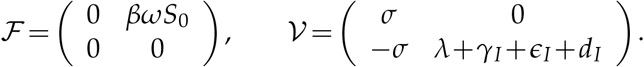

To evaluate the control of the disease timely, we introduce the corresponding effective control reproduction number as:

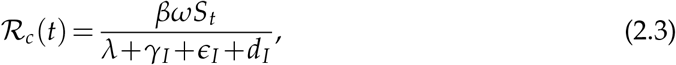

where *S*_*t*_ is the value of class *S* at time t. In the numerical simulation section, the tracking of the COVID-19 in Algeria can be clearly found and we will show the average effective control reproduction number in each stage.

## 3 Data

### 3.1 Preparation

In order to reflect the actual situation of the COVID-19 in Algeria, we adopt the combination of data and the model. Numerical simulation will estimate the values of some parameters of the epidemic model and the corresponding index will be given too. So, we can describe the disease quantitatively.

The daily new reported cases can be found in the website, but the primary health system and daily report mechanism are not excellent in Algeria. Especially, at the beginning of the pandemic, the nucleic acid detection capability is seriously inadequate and the experience of the respond to public health emergencies is insufficient. These factors are the main reasons that the epidemic situation reports exist delay and deficiency. There are some problems of the daily reported data, some data are vanished unexpected. In the initial collection, the new confirmed cases of some days are 0, because the spread of the disease is a continuous dynamic process, it is impossible. In order to avoid the negative effect of outliers, we will use the accumulated confirmed cases of COVID-19 in Algeria for simulation, which can benefit the numerical results intelligently. The source of the accumulated confirmed cases and COVID-19 induced death are from Algeria source [30–32] and Baidu [33].

### 3.2 Simulation procedure

After the outbreak of the COVID-19 in Algeria, the government pushed through a series of control strategies to prevent the disease from spreading, such as close all markets, all places of worship and parties, prohibited all family gatherings from 17 Mar. 2020. Though the aggregation activities have been forbidden for a long time, the people can still go out, they are not at home compulsively. Here we divide the simulation procedure into six periods according to the different control strategies.

I. 24 Feb. – 17 Mar. There is no strict policy.
II. 18 Mar. – 07 Apr. The first three weeks after carrying out the relatively strict policy (partial confinement strategy).
III. 08 Apr. – 12. May. The following six weeks after carrying out the relatively strict policy.
IV. 13 May – 13 Aug. Resumption to normal life gradually.
V. 14 Aug. – 25 Oct. Reopen the country ulteriorly.
VI. 26 Oct. – 30 Nov. Government requires inhabitants to maintain vigilant against the COVID-19.
VII. In the first stage, there is no strict control strategy to prevent the disease spread, so the spread of the COVID-19 at that time is under the natural state. Then, we use the estimated parameter *β* in the first stage as the value of contact rate through all the six stages. And in each stage, the recovery rates are updated according to the data fitting results.

### 3.3 Diagnostic Shadow

The spread of COVID-19 is really strong and its death rate is much higher than normal infectious diseases. There are many reports show that, the COVID-19 can spread asymptomatically and the symptoms of quite a few people are relatively mild [34, 35]. It brings many difficulties to follow all the infected people. So the number of the reported cases has been regarded as incompleted. Considering the data we collect, the crude mortality of the COVID-19 in Algeria is much higher than other countries, we conclude that the diagnostic shadow exists in the diagnosis of COVID-19 in Algeria.

To reflect the actual epidemic in Algeria, we introduce the method of diagnostic shadow into the data processing. China is considered to carry the most strict control strategy country, the government carry out free medical service for COVID-19 in the scope of further reducing the financial burden of the patients. And track everyone’s position to check the potential infected ones. These policies are made to ensure maximum the admission and cure of patients. Here, we define the statistical data in China as the standard mortality of COVID-19, it is known as 2.1% [36].

We define a time-dependent function *f* (*t*) to estimate the diagnostic shadow from the following identity,

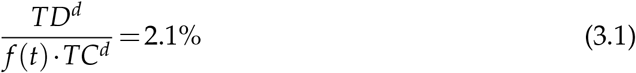

where *TC*^*d*^ and *TD*^*d*^ denote the accumulated confirmed and death cases, respectively, they are reported daily. Due to the oscillation of the daily reported data, we update the shadow value every three days (see Figure. 2).^†^ We calculated the average diagnostic shadow in each stage and listed in Table 1. Then, we estimated the accumulative confirmed population 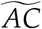 as

**Table 1:**
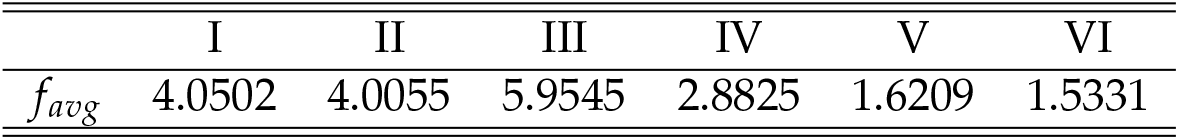
Average diagnostic shadow

**Figure 1:**
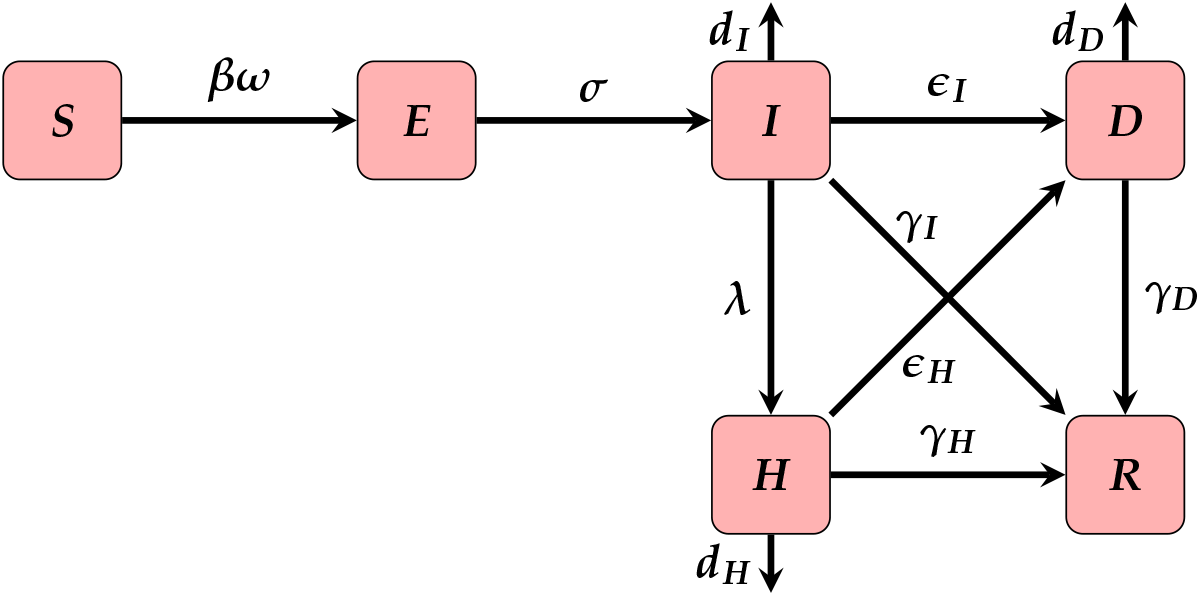
Illustration of the dynamical model

**Figure 2:**
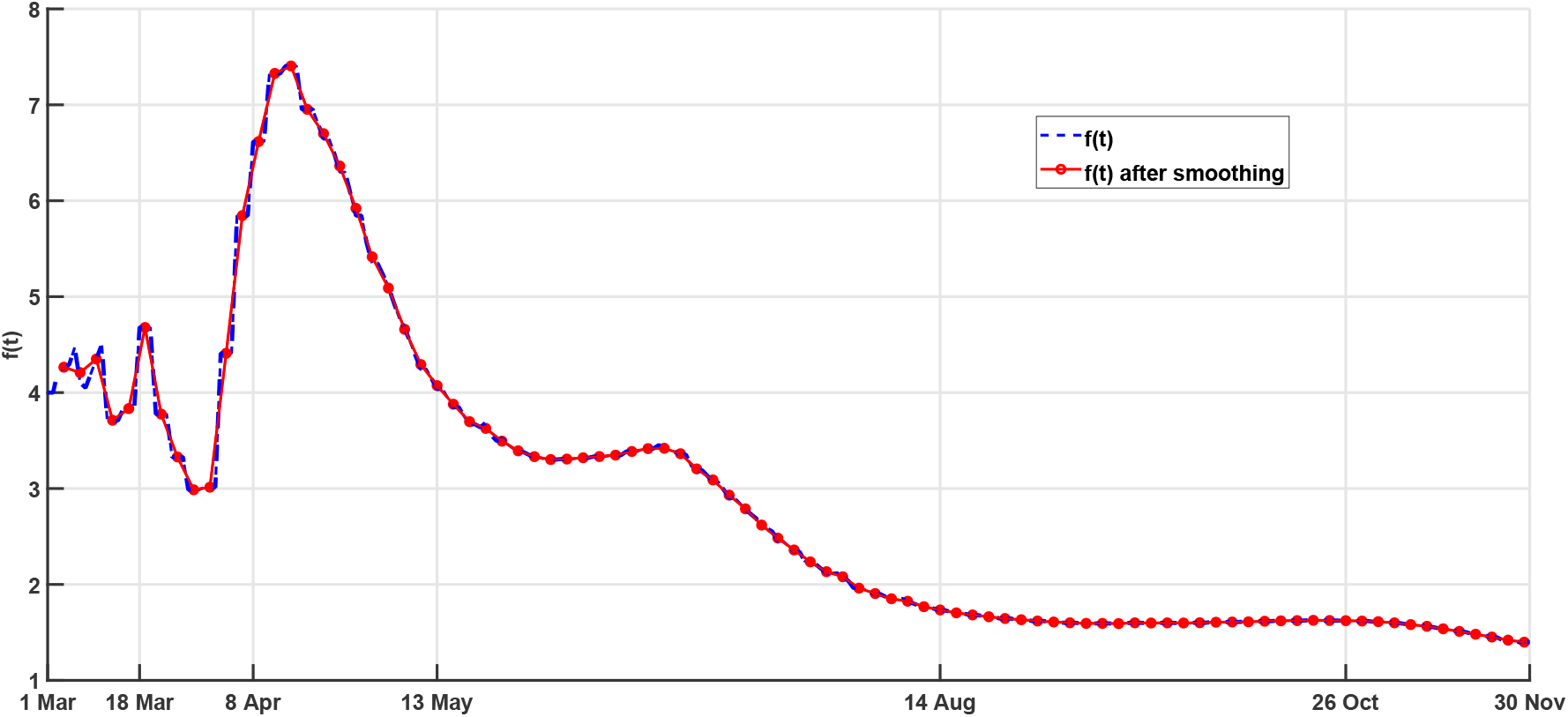
Diagnostic Shadow

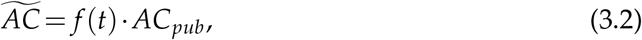

where *AC*_*pub*_ is the officially published accumulated infected population by Algeria government. In Figure. 2), at the beginning of the outbreak, the identification ability of the COVID-19 is insufficient, the average rate diagnostic shadow in first and second stage are almost the same. As the detection capability and medical treatment improved, it causes the rate of diagnostic shadow increased in the third stage and then decreased in the following stages. The diagnostic shadow also reflects the control effective level in Algeria and it has improved a lot. In the numerical simulation section, we use the processed data for fitting procedure. The well-handled data can reflect the epidemic more realistically.

## 4 Numerical simulation

### 4.1 Optimal data fit and tendency analysis

In model (2.1), we use the following formula to present the accumulated confirmed cases (*AC*) in period [*t*_0_,*t*],

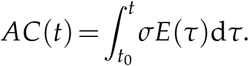

To overcome the limitation of the collected confirmed data, we take *AC*(*t*) as the variable for data fitting. The data which will be simulated is pre-processed as mentioned and the unit of the base time is one day. With the development of COVID-19 research, we have known more characteristics about the disease. Based on the clinical and statistics knowledge, we have the following assumptions and estimations on the parameters. Because there is no distinguish between the symptom and asymptomatic patients, the average incubation period (1/*σ*) we take here is longer than we did in the previous study. At the first two stage, it is taken as 21 days. And in the rest parts of the simulation, it is taken as 60 days. It is mainly because of the climate, high temperature will slow the spread of the diseases at some level [29]. As the pandemic developing, the awareness of people is changing with time. At the fist several stages, education and publicity play an important role in improving the awareness of people. It is reflected on the response of self-home quarantine. But in the later period, the corresponding assessment suggest the country can be reopen, the control strategy is slack after that. So we take the average self-home quarantine period (1/*λ*) from one to four weeks. The clinical diagnosis is limited to the ability of nucleic acid testing and basic medical security, we estimate the mean confirmed cycle of infected people (1/*ϵ*_*I*_) as 14 or 21 days and the mean period of self-home quarantine people turn to hospital treatment (1/*ϵ*_*H*_) as 7, 14 or 21 days according to different control stage. Using parameters *d*_*D*_ and *γ*_*D*_ in model (2.1), the disease mortality rate (*mr*) can be written as

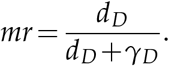

As mentioned above, we set *mr* as 2.1% for Algeria.

The values of parameter *γ*_*I*_ and *γ*_*H*_ are considered as the same. The patients will recover quicker under hospital treatment compared with the self-recovered population, so we fix the weight as *γ*_*D*_ = 1.1*γ*_*I*_. In the first stage, we think that, it is the nature contact rate (*β*_0_) which is considered to be the standard. The parameter *ω* reflects the change with time in different stage. All the mentioned parameter values are listed in Table 2 The total population size for optimal simulation is 4.39×10^7^ [30].

**Table 2:** Simulation parameter

| Parameter | I | II | III | IV | V | VI |
| --- | --- | --- | --- | --- | --- | --- |
| $\beta_0\omega$ | 4.0114E-08 | 4.8806E-08 | 1.0221E-09 | 5.6308E-09 | 1.2529E-09 | 1.9693E-08 |
| $\omega$ | 1 | 1.2167 | 0.0255 | 0.1404 | 0.0312 | 0.4909 |
| $\sigma$ | 1/21 | 1/21 | 1/60 | 1/60 | 1/60 | 1/60 |
| $\lambda$ | 1/28 | 1/14 | 1/7 | 1/7 | 1/21 | 1/21 |
| $\epsilon_I$ | 1/21 | 1/21 | 1/14 | 1/21 | 1/21 | 1/21 |
| $\epsilon_H$ | 1/14 | 1/14 | 1/7 | 1/21 | 1/21 | 1/21 |
| $\gamma_I$ | 0.1429 | 0.1419 | 0.1429 | 0.0100 | 0.0100 | 0.1429 |
| $\gamma_H$ | 0.1429 | 0.1419 | 0.1429 | 0.0100 | 0.0100 | 0.1429 |
| $\gamma_D$ | 0.1572 | 0.1561 | 0.1572 | 0.0110 | 0.0110 | 0.1572 |
| $d_I$ | 0.0045 | 0.0045 | 0.0045 | 0.0003 | 0.0003 | 0.0045 |
| $d_H$ | 0.0045 | 0.0045 | 0.0045 | 0.0003 | 0.0003 | 0.0045 |
| $d_D$ | 0.0034 | 0.0033 | 0.0034 | 0.0002 | 0.0002 | 0.0034 |

We employ a Least-Squares procedure to estimate parameters *β* and *γ*_*I*_ for each stage. Assuming that we have a proper estimations for other parameters in model (2.1), we need to solve the following optimization problem.

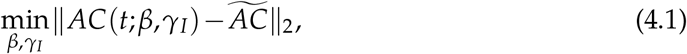

where 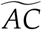 is the estimated infected population defined in equation (3.2). Then the estimation and prediction procedure are as follows.

Step 1 Set the initial condition {*S*(*t*_0_),*E*(*t*_0_), *I*(*t*_0_), *D*(*t*_0_), *H*(*t*_0_), *R*(*t*_0_)} and the proper values for the parameters in model (2.1) other than *β* and *γ*_*I*_.

Step 2 Based on the officially published data *AC*_*pub*_, calculate 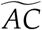 and solve the optimization problem (4.1) to obtain the estimations *β*^*^ and 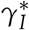 for *β* and *γ*_*I*_, respectively.

Step 3 Based on *β*^*^ and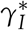, the initial condition and parameters set in Step 1, solve the dynamical system (2.1) to obtain *S*(*t*),*E*(*t*), *I*(*t*), *D*(*t*), *H*(*t*) and *R*(*t*).

The corresponding optimal simulations for each stage are presented in Figure 3. For the first stage, the time period is from 24 Feb. 2020 to 17 Mar. 2020 in which the disease hasn’t attract enough attention. By fitting model (2.1) to the accumulated confirmed COVID-19 cases with diagnostic shadow, we obtain the estimation for the transmission rate and denote it as *β*_0_. The Algeria government has announced that, all the place of worships, markets and gathering places must be closed, meanwhile, the family reunion is forbidden from 17 Mar. 2020. The second stage of our study is the first three weeks after carrying the relatively strict control policy. Due to the inertia of the transmission, the pandemic in Algeria at that time is still serious. It reflects on the *ω* value, it is greater than 1. From the optimal simulation results in Figure 3, Stage I and Stage II are both in the quick increase period. We show the total tendency of the COVID-19 development in Algeria in Figure 4, the yellow and red dotted lines represent the further trends in Algeria if the former control policy keeps on, respectively. It is obvious that, at that two periods, the disease will be out of control. In a more quantized manner to show the results, we calculate the average value of the effective control reproduction number for each stage and show them in Table 3. The basic reproduction number of Algeria is estimated as 7.63 which is a little higher than that in China [19]. From Table 3, the value of ℛ_*c*_(*t*) in Stage I is greater than Stage II.

**Table 3:** Average *R*_*c*_ (*t*) for each stage

|  | I | II | III | IV | V | VI |
| --- | --- | --- | --- | --- | --- | --- |
| $R_c(t)$ | 7.6300 | 8.0703 | 0.1239 | 1.2291 | 0.5199 | 3.5529 |

**Figure 3:**
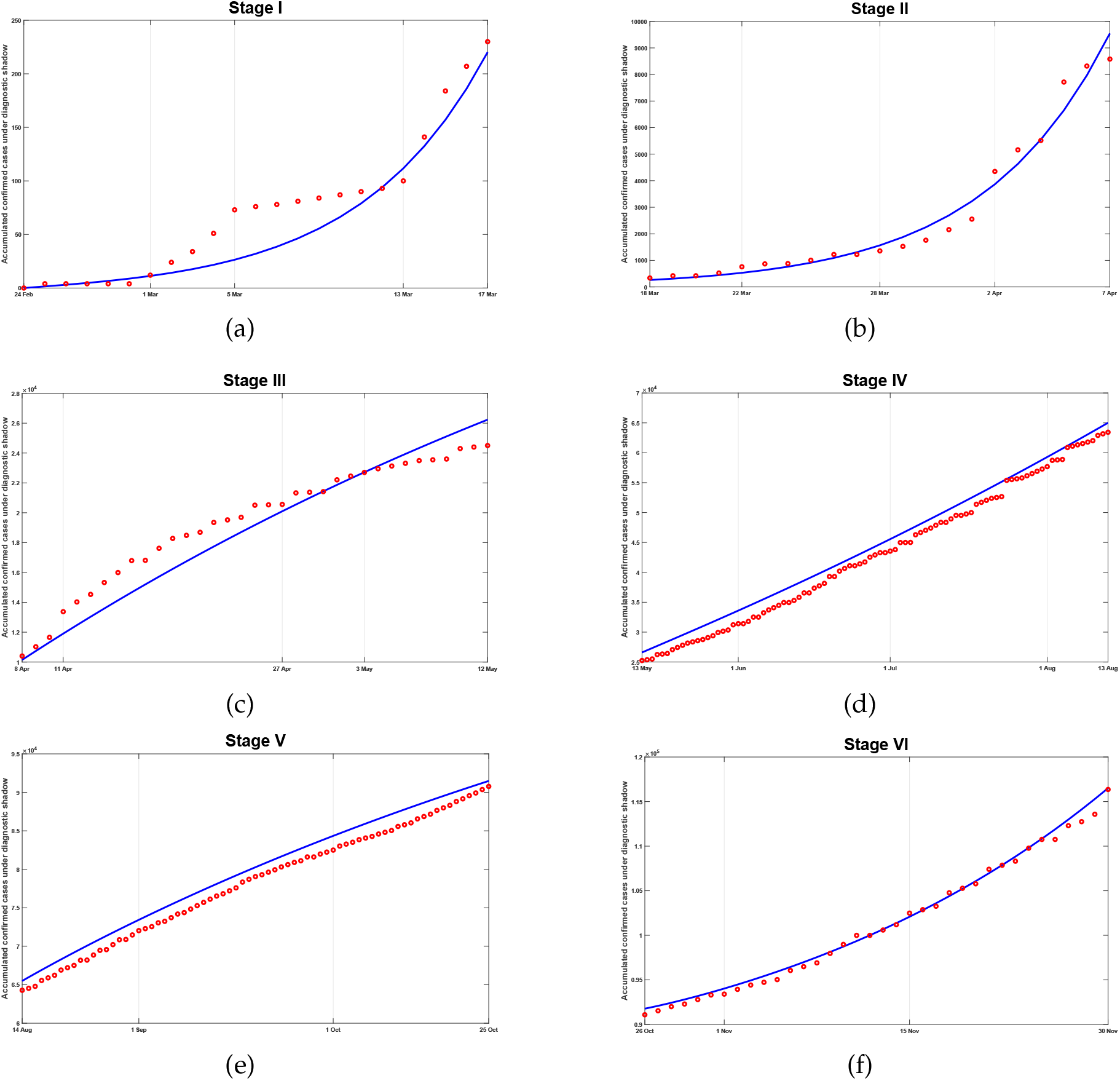
Accumulated confirmed cases simulation for each stage. Red circle: 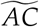; Blue solid line: *AC*(*t*).

**Figure 4:**
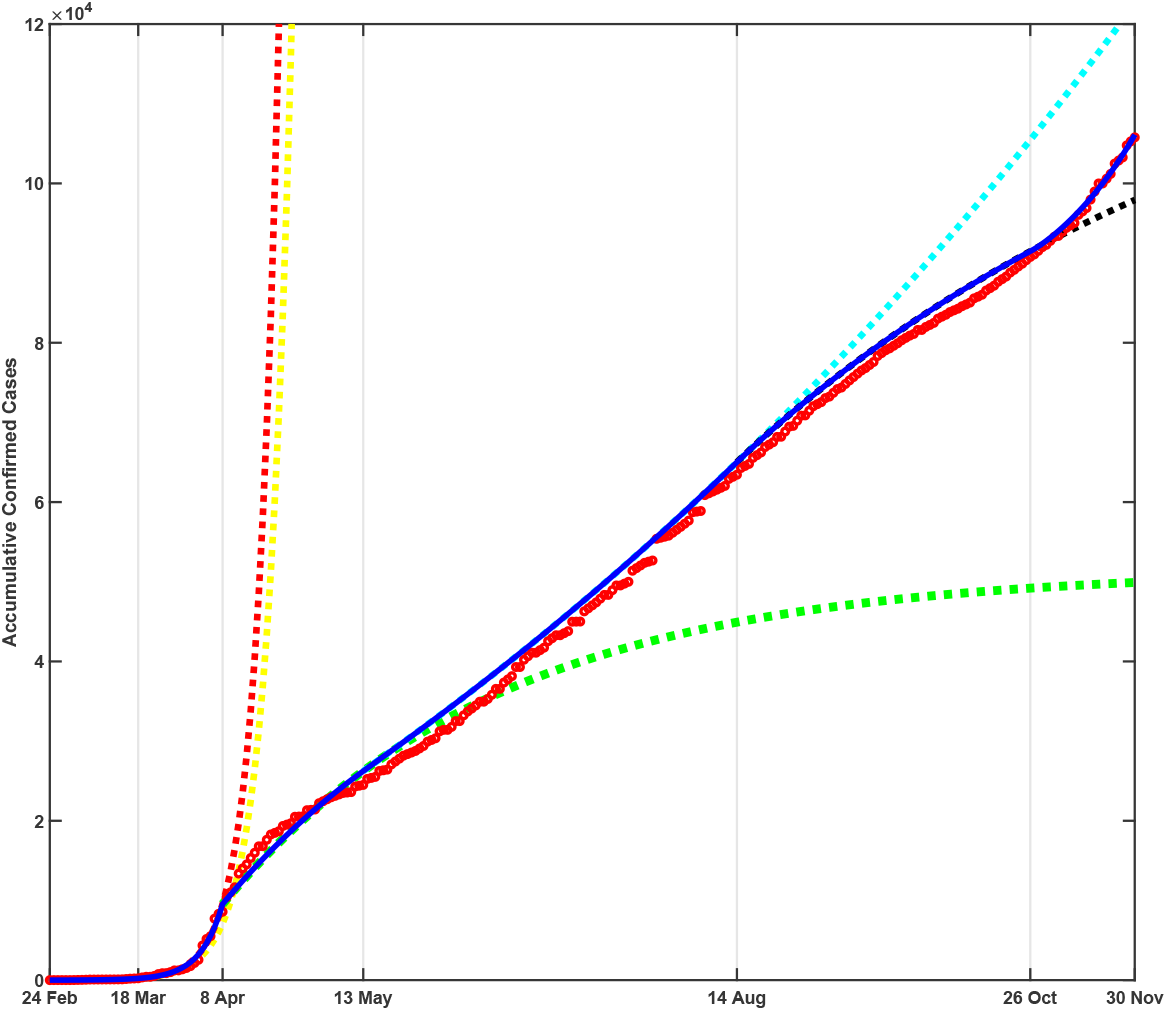
Simulation result. Red circle: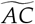; Blue solid line: *AC*(*t*). Dotted lines: simulation result for keeping the control strategy from Stage I(yellow), II(red), III(green), IV(blue-green) and V(black).

In the third stage, the effect of the relatively strict isolation strategy can be seen clearly in Figure 3(c). The growth of accumulated confirmed cases is slowing down and the effective control reproduction number in that time is 0.1239 which is the smallest among the six stages and is much smaller than 1. If the government can consist the control strategy like this, the final size of accumulative confirmed cases will be much smaller than the actual situation and we show this numerical result with green dotted line in Figure 4. But it is theoretically that the whole country can persist relatively strict isolation strategy for such a long time, normalized prevention and control is an inexorable trend. As the weather becomes warmer and the epidemic of COVID-19 tends to a good prospect, Algeria government decide to resume normal life gradually. The effective reproduction number of Stage IV is greater than 1 again. It is the transition from relatively strict control strategy to normalized prevention. If the normalized control strategy cannot be proceeding well, just like the fourth stage in Algeria, the accumulated confirmed cases will be increased further which we have shown with blue-green dotted line in Figure 4. After the transitory stage, on 14 Aug. 2020, the country reopen ulteriorly. Although the effective reproduction number of Stage IV is 0.5199(< 1) which is greater than Stage III, it is enough for daily disease prevention. We can see the obvious progress of the epidemic control in Algeria with balck dotted line in Figure 4.

The COVID-19 has a distinct seasonal characteristics. In autumn and winter, the viral infection rate is high. The cross infection of flu and COVID-19 makes the situation more complex. The pandemic caused by COVID-19 resurgence around the world. From 26 Oct. 2020, Algeria government urges the citizen keep vigilant against the COVID-19. But without the relatively strict isolation strategy, the disease is going around again in Algeria, the effective control reproduction number of Stage VI is estimated by optimal simulation as 3.5529 which is greater than 1. And in Figure 3(f), from the convexity of the curve, it has a great possible to cause a massive outbreak. The economic in Algeria has shown recession historically. Lockdown strategy may not be the best way to disease control. There are some good news about the vaccines for the COVID-19, in the next control period, we may have to pin our hope on vaccination strategy and specific medicine.

### 4.2 Medical Resource Estimation

In this part, we will discuss the medical resource in Algeria. There are two types of health structures and six types of health workers in Algeria, we summarize the descriptions and abbreviations of the structures and workers in Table 4. MR-related data are listed in Table 5.

**Table 4:** Descriptions and abbreviations

| The public health structures |  |  |  |
| --- | --- | --- | --- |
| BHU | Basic health units | HSF | Hospital facilities |
| BHC | Basic health care structures | HCT | Health centers |
| GHS | General hospitals | UHS | University hospitals |
| SHS | Specialized hospitals |  |  |
| The private health structures |  |  |  |
| SMC | Surgical medical clinics | MCL | Medical clinics |
| BCC | Blood clearing centers | RAC | Reproductive assistance centers |
| HTU | Health transport units | SCC | Specialized consulting clinics |
| GCC | General consultation clinics |  |  |
| The health workers |  |  |  |
| APB | Active in the public sector | APR | Active in the private sector |
| PMA | Paramedical aids | SDH | State degree holders |
| QUL | Qualifications | PAS | Paramedical assistants |

**Table 5:**
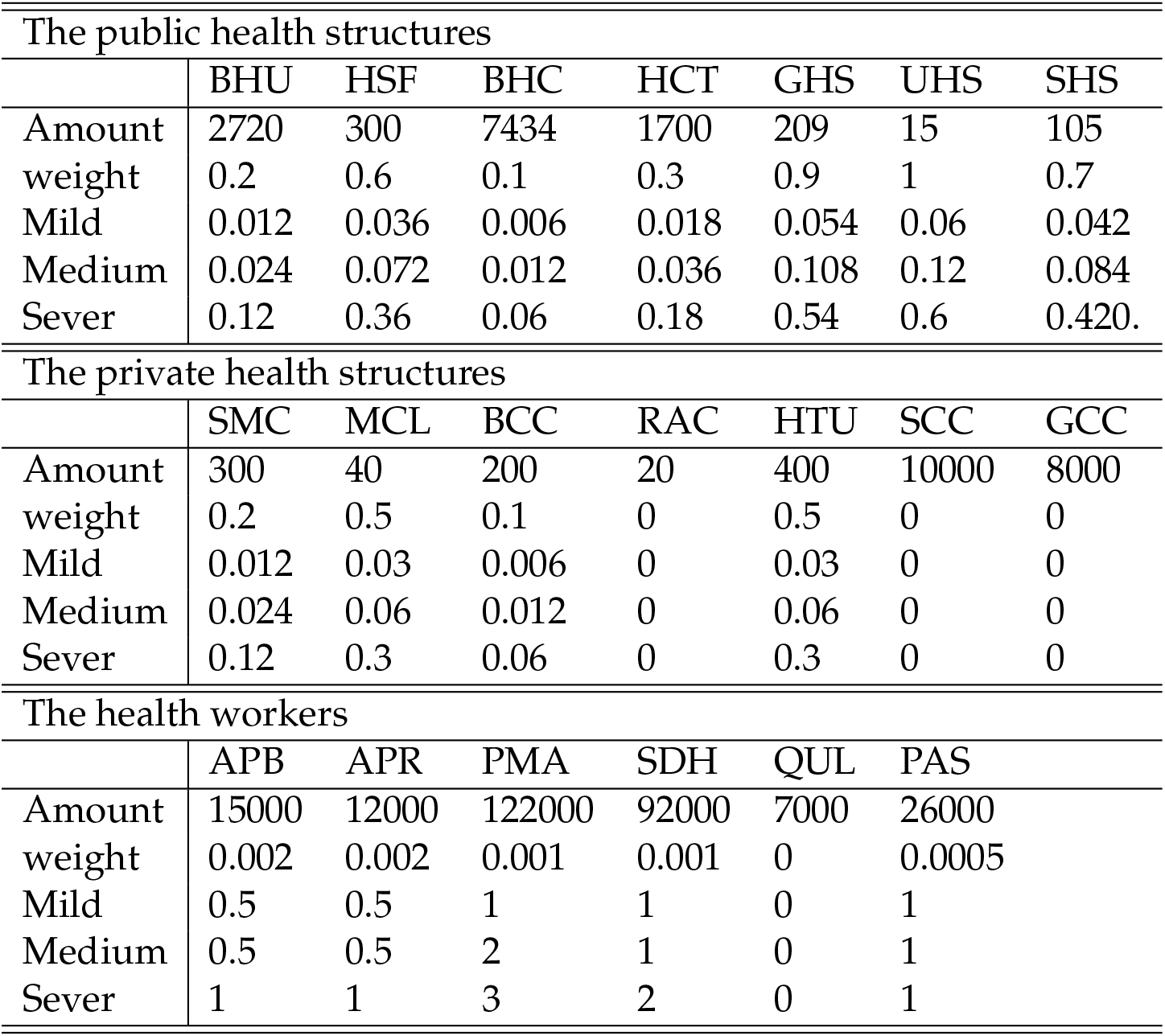
Medical resource estimation

| The public health structures |  |  |  |  |  |  |  |
| --- | --- | --- | --- | --- | --- | --- | --- |
|  | BHU | HSF | BHC | HCT | GHS | UHS | SHS |
| Amount | 2720 | 300 | 7434 | 1700 | 209 | 15 | 105 |
| weight | 0.2 | 0.6 | 0.1 | 0.3 | 0.9 | 1 | 0.7 |
| Mild | 0.012 | 0.036 | 0.006 | 0.018 | 0.054 | 0.06 | 0.042 |
| Medium | 0.024 | 0.072 | 0.012 | 0.036 | 0.108 | 0.12 | 0.084 |
| Sever | 0.12 | 0.36 | 0.06 | 0.18 | 0.54 | 0.6 | 0.420. |
| The private health structures |  |  |  |  |  |  |  |
|  | SMC | MCL | BCC | RAC | HTU | SCC | GCC |
| Amount | 300 | 40 | 200 | 20 | 400 | 10000 | 8000 |
| weight | 0.2 | 0.5 | 0.1 | 0 | 0.5 | 0 | 0 |
| Mild | 0.012 | 0.03 | 0.006 | 0 | 0.03 | 0 | 0 |
| Medium | 0.024 | 0.06 | 0.012 | 0 | 0.06 | 0 | 0 |
| Sever | 0.12 | 0.3 | 0.06 | 0 | 0.3 | 0 | 0 |
| The health workers |  |  |  |  |  |  |  |
|  | APB | APR | PMA | SDH | QUL | PAS |  |
| Amount | 15000 | 12000 | 122000 | 92000 | 7000 | 26000 |  |
| weight | 0.002 | 0.002 | 0.001 | 0.001 | 0 | 0.0005 |  |
| Mild | 0.5 | 0.5 | 1 | 1 | 0 | 1 |  |
| Medium | 0.5 | 0.5 | 2 | 1 | 0 | 1 |  |
| Sever | 1 | 1 | 3 | 2 | 0 | 1 |  |

1. The amount (2nd line, Table 5) of each type of health structures and workers are collected from the official website of Algerian Ministry of Health [30].
2. The weight of daily MR capacity per unit (3rd line, Table 5) and the daily MR costed (4th-6th lines, Table 5) by different types (Mild, Medium, Sever) of patients are estimated.
3. The baseline of daily MR capacity (*MR*_*base*_) in Algeria is calculated by multiplying Amount and *h*· weight in Table 5, *h* = 10^6^ is a calibrating coefficient. *MR*_*base*_ is displayed in Figure 5 by red dashed line.
4. The daily MR capacity varies with the development of the epidemic, we take the average diagnostic shadow (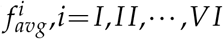, presented in Table 1) as an indicator to estimate the variation of daily MR capacity for each stage. We use 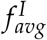 as the datum, *MR*_*base*_ as the medical resource in Stage I, and

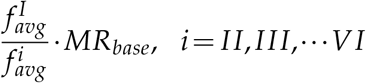

as the medical resource for other stages, which are plotted in Figure 5 by color dotted lines.
5. The proportion of the daily hospitalization patients *D*(*t*) for each stage are estimated in Table 6.
6. The daily demanded MR is calculated by multiplying the proportion in Table 6, the daily hospitalization patients *D*(*t*), the amount and the daily MR costed in Table 5, which are presented in Figure 5 by color solid lines.

**Figure 5:**
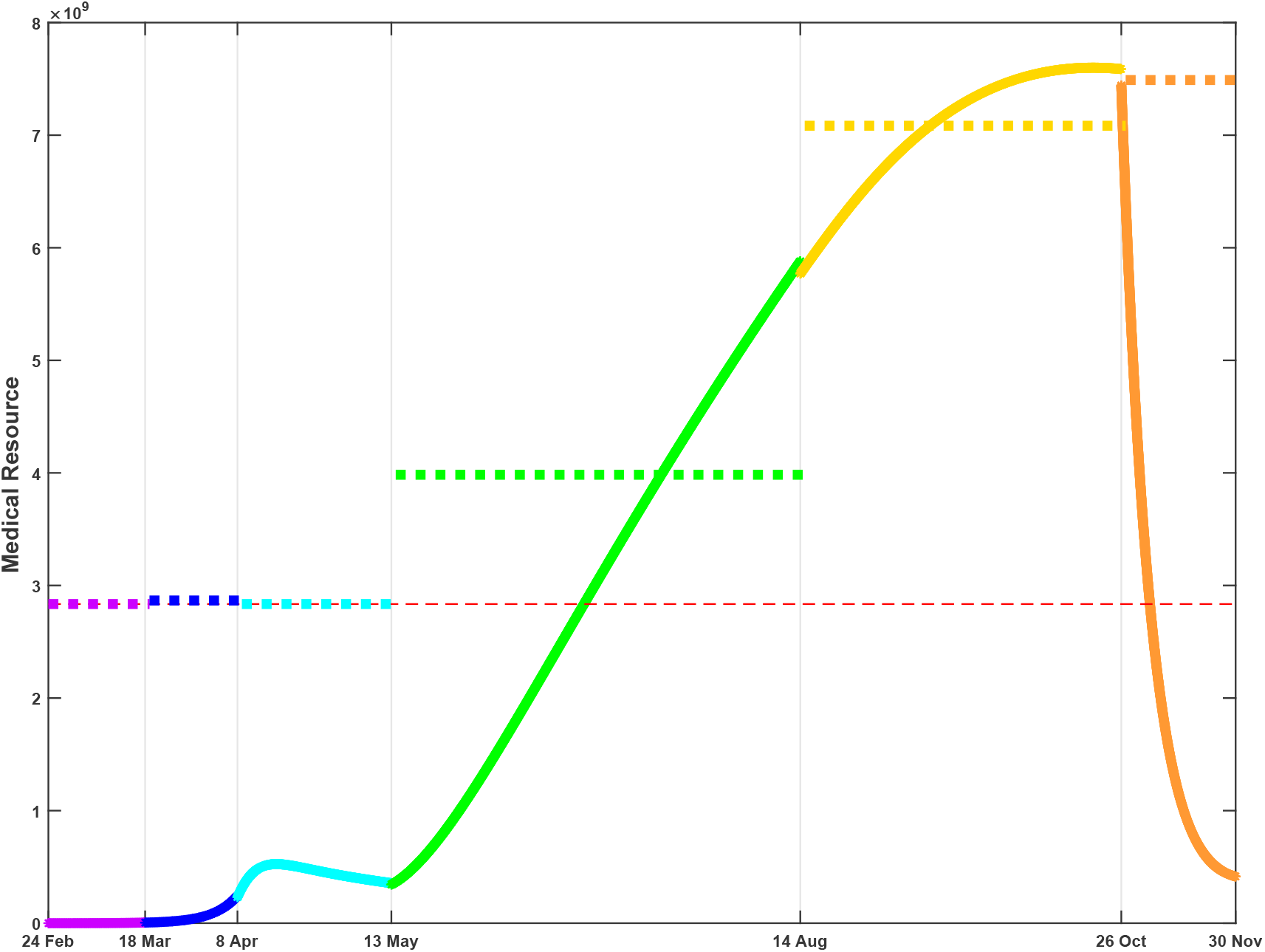
Medical Resource (MR) Estimation. Solid line: the daily demanded MR; dotted line: the daily MR capacity; dashed line: MR baseline.

**Table 6:** Estimation of the proportion of patients with different symptoms

| % | I | II | III | IV | V | VI |
| --- | --- | --- | --- | --- | --- | --- |
| Mild | 40 | 45 | 50 | 60 | 65 | 70 |
| Medium | 40 | 40 | 40 | 30 | 25 | 20 |
| Sever | 20 | 15 | 10 | 10 | 10 | 10 |

Figure 5 compares the daily demanded MR and the estimated daily MR capacity. At the beginning, the medical resource covered the requirement well; with the rapid development of the epidemic, the daily demand overflowed in the fourth and fifth stages. At the same time, the daily MR capacity was growing with the experience and the international assistance. The daily demand declined underneath the warning line in the sixth stage. Adequate medical resource is crucial for controlling the epidemic, many countries suffer from the medical burden caused by COVID-19 [42–44]. Designated and mobile cabin hospitals have been playing a vital role in the prevention of disease spreading [19]. To ensure the medical treatment, China government sets up many designated hospitals for each province and mobile cabin hospitals for provinces with sever epidemic, which released the medical burden sharply and provided earlier diagnosis, earlier isolation and better treatment service [45, 46]. And this experience has been generalized to many other countries.

## 5 Conclusion

An SEIR-based dynamical model is founded to describe the COVID-19 development in Algeria. Base on the control strategy, the simulation procedure is divided into six stages with different parameter settings. Due to the abnormal mortality rate in Algeria, a time-dependent diagnostic shadow is defined to estimate the accumulated confirmed cases. The proposed diagnostic shadow *f* (*t*) plays an important role in the numerical simulation procedure. The effective reproduction number ℛ_*c*_(*t*) is calculated for each stage. It shows that the epidemic is growing fast in Stage I and II, with ℛ_*c*_(*t*) much greater than 1. In Stage III, Algeria government performs the relatively strict isolation strategy, ℛ_*c*_(*t*) drops sharply. After the resumption to normal life in Stage IV, V and VI, ℛ_*c*_(*t*) increases oscillated with the rigorousness of the strategy. The simulation results show the effectiveness of the isolation strategy performs in Stage III, if such strategy keeps until the end of Nov. 2020, the accumulated confirmed cases should be halved. Both capacity and demanded medical resources are also estimated, with the rapid development of the accumulated confirmed cases, the demanded medical resource overwhelmed the capacity in Stage IV and V, and the medical burden release in Stage VI. This retrospective analysis suggests that proper isolation is still an effective prevention strategy from the COVID-19 rapid spreading.

## Data Availability

The data used in this study are all public data. Contact the author if necessary.

## Acknowledgments

This work was partially supported by National Natural Science Foundation of China (Grant Nos. 41704116, 11901234), Jilin Provincial Excellent Youth Talents Foundation (Grant No. 20180520093JH), Scientific Research Project of Education Department of Jilin Province (Grant No. JJKH20200933KJ), Fundamental Research Funds for the Central Universities, JLU (Grant No. 93K172020K27).

## Footnotes

† Due to lacking of the data for the first a few days, we estimated the diagnostic shadow from 1 Mar., 2020.

